# Association Between Primary Tumour–Brain Metastasis Receptor Status Mismatch and Outcomes in Breast Cancer Brain Metastasis

**DOI:** 10.1101/2023.01.18.23284724

**Authors:** Yilong Zheng, Chen Ee Low, Sheryl Yen Pin Tan, Clarisse Wei Yen Hing, Chun En Yau, Kejia Teo, Vincent Diong Weng Nga, Tseng Tsai Yeo, Andrea Li-Ann Wong, Mervyn Jun Rui Lim

**Affiliations:** Yong Loo Lin School of Medicine, National University of Singapore, Singapore; Division of Neurosurgery, National University Hospital, Singapore; Department of Haematology-Oncology, National University Cancer Institute, Singapore

**Author notes:** Co-first authors (Yilong Zheng, Chen Ee Low). **<u>Details of the Corresponding Author</u>** Yilong Zheng, Yong Loo Lin School of Medicine, National University of Singapore, Singapore, 10 Medical Drive, Singapore 117597.

**Keywords:** biomarker, risk stratification, brain cancer

## Abstract

**Objective:** To evaluate the association between primary tumour–brain metastasis receptor status mismatch and outcomes in patients with breast cancer brain metastasis (BCBM).

**Methods:** Patients who (1) had a histologically verified breast-to-brain metastasis, (2) were 18 years old or above on the day of surgical resection, and (3) had the ER (estrogen receptor), PR (progesterone receptor), and HER2 (human epidermal growth factor receptor 2) statuses of both the primary breast tumour and the secondary brain metastasis were retrospectively recruited. Univariate time-to-event analysis was performed using the Kaplan-Meier method. The exposures analysed were the various combinations of ER, PR, and HER2 statuses between the primary breast tumour and the secondary brain metastasis. The outcomes were overall mortality and recurrence.

**Results:** Of the 158 patients who underwent surgical resection of brain metastases during the study period, 31 were included in the analysis. The mean (SD) age of the study population was 56.7 (12.2), and most patients were Chinese (54.8%). On univariate analysis of the association between the various receptor status combinations and overall mortality, ER (p=0.920) and PR (p=0.390) status conversion were both found to not be associated with overall mortality. However, HER2 status conversion was found to be significantly associated with overall mortality (p=0.026). Specifically, patients whose primary tumour was HER2+ but whose secondary brain metastasis was HER2− had the poorest outcome, with a median overall survival of 3.4 months. On the other hand, the median overall survival of the other HER2 receptor status combinations ranged from 10.9 to 16.6 months. There were no statistically significant associations between status conversion of any of the receptors and recurrence.

**Conclusions:** Among patients who underwent surgical resection of BCBMs, patients with primary tumour HER2+ but secondary brain metastasis HER2− had a significantly higher risk of mortality. However, ER and PR status conversion was not significantly associated with outcomes.

## Introduction

Brain metastases are the most common intracranial tumors in adults (1), and the breast is the second most common primary cancer site (2). With modern healthcare along with better treatment options and longer lifespans of breast cancer patients, the incidence of breast cancer brain metastasis (BCBM) has increased (3, 4).

BCBM carries a poor prognosis with existing treatment options, with median survival generally spanning 2–9 months (5, 6). Available treatment options primarily include local therapies such as surgical resection, radiotherapy, and/or stereotactic radiosurgery. Systemic therapies such as chemotherapy and targeted therapy generally have limited impact on outcomes (7), mainly because of the low blood-brain barrier penetrance of these agents. Novel systemic management for patients unsuitable for local therapies include systemic therapies which can cross the blood-brain barrier, such as targeted small-molecule tyrosine kinase inhibitors, and combinations with pre-existing agents (8).

To maximise the therapeutic activity of systemic therapy through individualized management, tumour-intrinsic factors, such as tumour hormone and human epidermal growth factor receptor 2 (HER2) receptor status, must be considered. Recognizing receptor statuses associated with brain metastasis and propagation can thus aid with risk stratification, predict efficacy of systemic therapy, allow for personalized therapy, and alter clinical trajectory for BCBM patients.

However, prior studies have established receptor conversions in up to 20-50% of BCBM patients (9, 10), potentially reducing the viability of such personalised therapies. In such cases where the receptor status between the primary tumour and the brain metastasis differ, prognosis is uncertain, and the efficacy of individualized systemic therapy may also differ. In this study, we aimed to demonstrate and discuss the correlation between receptor conversion in BCBM and overall mortality and recurrence.

## Methodology

### Study design

This was a retrospective study of patients who underwent surgical resection of BCBMs at our institution between March 2011 and December 2019. Institutional ethics approval was obtained from the institutional review board prior to study initiation (National Healthcare Group Domain Specific Review Board; Reference Number 2020/00358).

### Cohort selection

The operating theatre records database was accessed to retrieve the list of patients who underwent surgical resection of brain metastases during the study period. The electronic medical records of the patients were then accessed, and the records were screened for patients who met the inclusion criteria. Patients who (1) had a histologically verified breast-to-brain metastasis, (2) were 18 years old or above on the day of surgical resection, and (3) had the ER, PR, and HER2 statuses of both the primary breast tumour and the secondary brain metastasis were included in the analysis.

### Exposures and outcomes

The exposures analysed in this study were the various combinations of ER, PR, and HER2 statuses between the primary breast tumour and the secondary brain metastasis. The outcomes were overall mortality and recurrence (as adjudicated by the radiologists).

### Statistical analysis

The baseline characteristics of the patients were reported using mean and standard deviation for continuous variables, count numbers and percentages for categorical variables. Univariate time-to-event analysis was performed using the Kaplan-Meier method. Hypothesis testing was performed using the log-rank method, and a p-value of lower than 0.050 was regarded as statistically significant. All data analyses were conducted using R Studio Version 1.2.5042.

## Results

### Baseline characteristics of the study population

Of the 158 patients who underwent surgical resection of brain metastases during the study period, 31 were included in the analysis. The baseline characteristics of the study population were reported in Table

1. The mean (SD) age of the study population was 56.7 (12.2), and most patients were Chinese (54.8%). Most patients did not have conversion of receptor statuses.

### Association between receptor conversion and outcomes

On univariate time-to-event analysis of the association between the various combinations of receptor statuses and overall mortality, ER (p=0.920) and PR (p=0.390) status conversion were both found to not be associated with overall mortality (Figure 1A and 1B). However, HER2 status conversion was found to be significantly associated with overall mortality (p=0.026) (Figure 1C). Specifically, patients whose primary tumour was HER2+ but whose secondary brain metastasis was HER2− had the poorest outcome, with a median overall survival of 3.4 months. On the other hand, the median overall survival of the other HER2 receptor status combinations ranged from 10.9 to 16.6 months. There was no statistically significant association between conversion of any of the receptors and recurrence (Figure 2).

## Discussion

In this study, we demonstrated that HER2 receptor conversion was significantly associated with overall mortality. Specifically, conversion from HER2+ to HER2− was associated with the poorest prognosis. However, ER and PR status conversion was not associated with either overall mortality or recurrence. These findings are concordant with the findings of other similar studies in the literature (11, 12).

We hypothesize that receptor conversion is a sign of genomic instability; specifically, the tumour genomes of patients with receptor conversion are likely to be more unstable than the genomes of patients without receptor conversion. As genomic instability is generally accepted to be associated with poorer outcomes (13), it follows logically that patients with receptor conversion would also have poorer outcomes.

However, while receptor conversion may be associated with poorer outcomes, some cases of conversion may be treatable with targeted therapy. For example, in cases where patients had primary HER2− but secondary HER2+, the conversion from HER2− to HER2+ can be managed with Herceptin. This could explain why patients with conversion from HER2− to HER2+ had similar outcomes with patients without HER2 receptor conversion (Figure 1C), despite having possibly greater genomic instability. On the other hand, there are limited treatment options for conversions from HER2+ to HER2−. Coupling this with the greater genomic instability and hence inherently poorer prognosis, patients with HER2+ to HER2− conversion have the poorest prognosis.

Interestingly, however, conversion of ER and PR status was not significantly associated with poorer outcomes. The reasons for this observation are unclear, and further studies are needed to explain this finding.

Several limitations of our study merit mention. First, the statistical validity of our findings may be limited, in view of the small sample size of our cohort. The small sample size of our cohort also precluded us from conducting multivariate analyses, in view of the possibility of model overfitting. Second, we analysed only data from our institution, and therefore our findings may not be generalisable to other institutions. Also, as only patients who underwent surgical resection of BCBMs were included in our analysis, our findings may not be generalisable to patients who did not undergo surgical resection of BCBMs.

## Conclusions

Among patients who underwent surgical resection of BCBMs, patients with primary tumour HER2+ but secondary brain metastasis HER2− had a significantly higher risk of mortality. However, ER and PR status conversion was not significantly associated with outcomes.

## Data Availability

All data produced in the present study are available upon reasonable request to the authors

## Acknowledgements

None.

