## Supplementary material for "Association Between Primary Tumour–Brain Metastasis Receptor Status Mismatch and Outcomes in Breast Cancer Brain Metastasis": .

Table 1: Baseline characteristics of the study population

| **Variable** | **Total (n=31)** |
| --- | --- |
| Age; mean (SD) | 56.7 (12.2) |
| Ethnicity; *n* (%) |  |
| Chinese | 17 (54.8) |
| Malay | 7 (22.6) |
| Indian | 4 (12.9) |
| Others | 3 (9.7) |
| Preoperative ECOG status; mean (SD) | 1.1 (0.6) |
| Number of brain metastases on preoperative MRI; *n* (%) | 2.1 (1.7) |
| Presence of extracranial metastases preoperatively; *n* (%) | 18 (58.1) |
| Presence of brain metastases by location; *n* (%) |  |
| Temporal | 7 (22.6) |
| Frontal | 8 (25.8) |
| Occipital | 7 (22.6) |
| Parietal | 11 (35.5) |
| Cerebellum | 16 (51.6) |
| Volume of largest brain metastasis; mean (SD) | 19 (13.5) |
| Extent of resection; *n* (%) |  |
| Gross total resection | 27 (87.1) |
| Subtotal resection | 4 (12.9) |
| Adjuvant therapy; *n* (%) |  |
| Radiotherapy | 6 (19.4) |
| Chemotherapy | 6 (19.4) |
| Gamma Knife radiosurgery | 7 (22.6) |
| Whole brain radiotherapy | 9 (29.0) |
| ER status; *n* (%) |  |
| Primary breast and secondary brain metastasis both negative | 14 (45.2) |
| Primary breast and secondary brain metastasis both positive | 12 (38.7) |
| Primary breast negative, secondary brain metastasis positive | 1 (3.2) |
| Primary breast positive, secondary brain metastasis negative | 4 (12.9) |
| PR status; *n* (%) |  |
| Primary breast and secondary brain metastasis both negative | 16 (51.6) |
| Primary breast and secondary brain metastasis both positive | 8 (25.8) |
| Primary breast negative, secondary brain metastasis positive | 5 (16.1) |
| Primary breast positive, secondary brain metastasis negative | 2 (6.5) |
| HER2 status; *n* (%) |  |
| Primary breast and secondary brain metastasis both negative | 12 (38.7) |
| Primary breast and secondary brain metastasis both positive | 14 (45.2) |
| Primary breast negative, secondary brain metastasis positive | 2 (6.5) |
| Primary breast positive, secondary brain metastasis negative | 3 (9.7) |

Abbreviations: SD, standard deviation; ECOG, Eastern Cooperative Oncology Group; MRI, magnetic resonance imaging; ER, estrogen receptor; PR, progesterone receptor; HER2, human epidermal growth factor receptor 2.

Figure 1: Kaplan-Meier curves evaluating the association between difference in hormone receptor status between the primary tumor and the resected brain metastasis and overall mortality. (A) ER status, (B) PR status, (C) HER2 status.

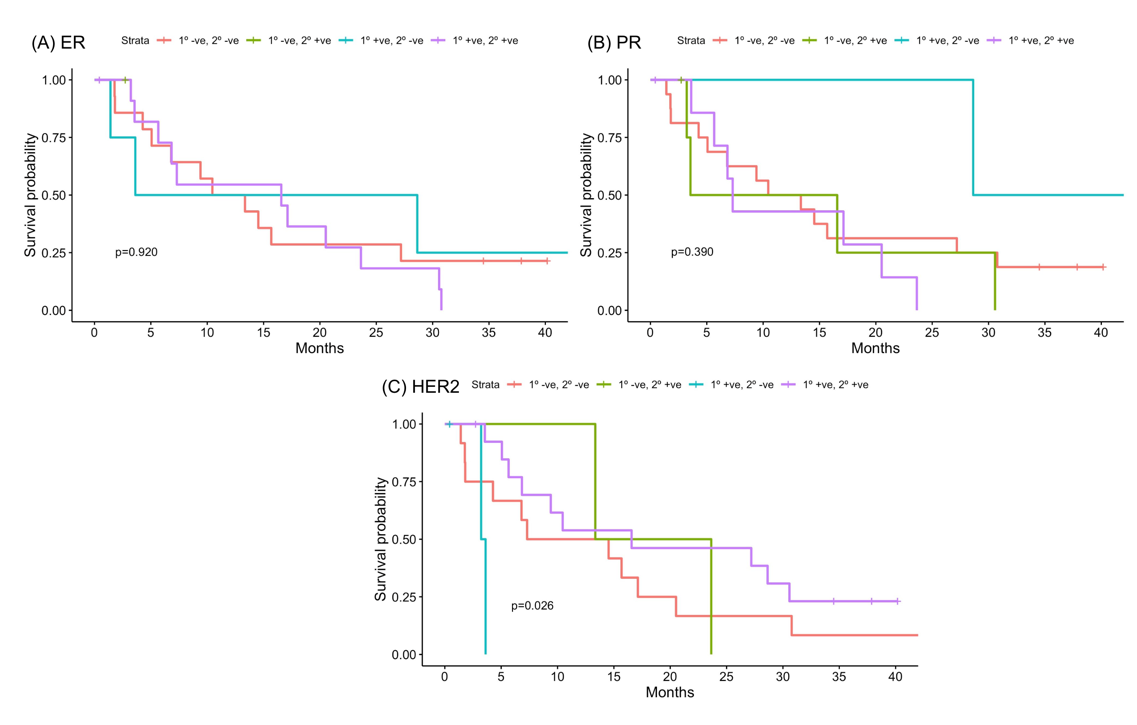

Abbreviations: ER, estrogen receptor; PR, progesterone receptor; HER2, human epidermal growth factor receptor 2; 1°, primary breast; 2°, secondary brain metastasis; +ve, positive; -ve, negative.

Figure 2: Kaplan-Meier curves evaluating the association between difference in hormone receptor status between the primary tumor and the resected brain metastasis and recurrence. (A) ER status, (B) PR status, (C) HER2 status.

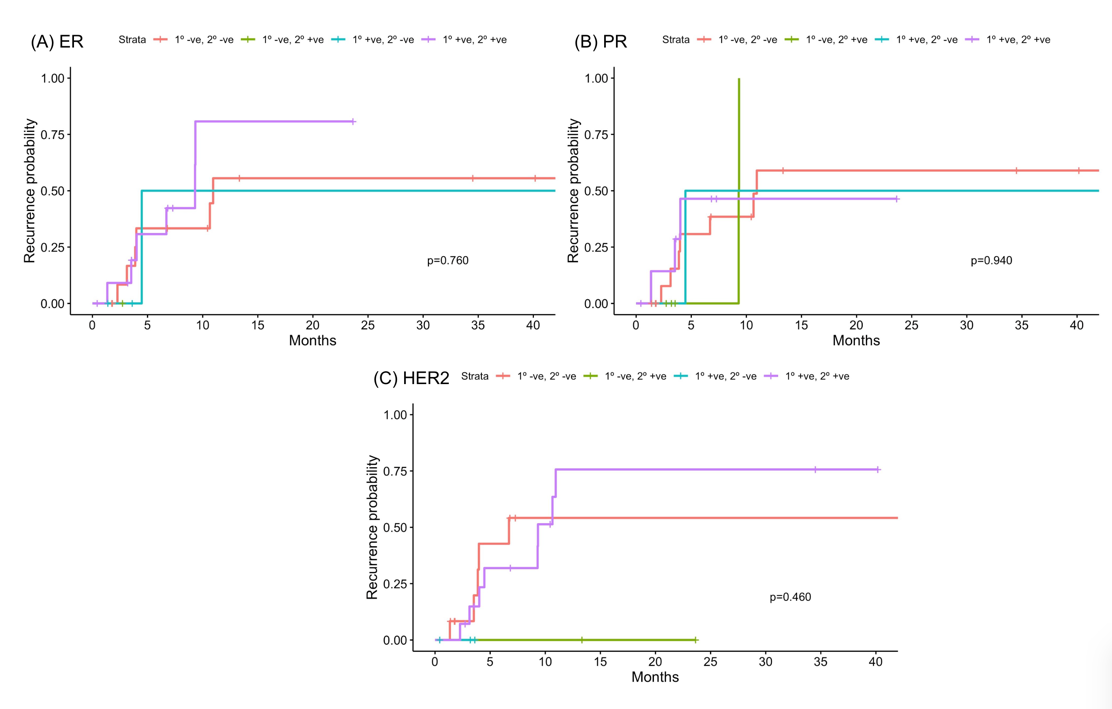

Abbreviations: ER, estrogen receptor; PR, progesterone receptor; HER2, human epidermal growth factor receptor 2; 1°, primary breast; 2°, secondary brain metastasis; +ve, positive; -ve, negative.
